# Individual cases of Parkinson’s disease can be robustly classified by cortical oscillatory activity from magnetoencephalography

**DOI:** 10.1101/2024.08.27.24312669

**Authors:** Gillian Roberts, Samuel Hardy, Robert Chen, Benjamin T Dunkley, PREVENT-AD Research Group & Quebec Parkinson Network

## Abstract

Parkinson’s disease (PD) is a progressive neurodegenerative disorder which causes debilitating symptoms in both the motor and cognitive domains. The neurophysiological markers of PD include ‘oscillopathies’ such as diffuse neural oscillatory slowing, dysregulated beta band activity, and changes in interhemispheric functional connectivity; however, the relative importance of these markers as determinants of disease status is not clear. In this study, we used resting state magnetoencephalography data (n = 199 participants, 78 PD, 121 controls) from the open OMEGA repository to investigate changes in spectral power and functional networks in PD. Using a Contrast of Parameter Estimates (COPE) approach, we modelled the effects of PD while controlling for population-level confounds (age, sex, brain volume). Permutation testing revealed highly significant increases in theta (p=0.0001) and decreases in gamma band spectral power (p=0.0001). Building on the group contrast results, we investigated the ability of source-resolved MEG data to distinguish PD from healthy controls. Our approach uses a Partial Least Squares (PLS)-based classifier to find linear combinations of MEG features which independently predict PD. We found MEG-based predictions to be highly sensitive and specific, reaching an optimal AUC-ROC of 0.87 ± 0.04 using a model including spectral power features with 4 independent PLS components, compared to 0.68 ± 0.04 when using functional connectivity. Interpretation of the model weights suggests that oscillatory slowing can be separated into independent posterior theta and global diffuse delta components that can robustly identify individual cases of PD with a high degree of accuracy. This suggests MEG can reveal dissociable, complementary neural processes which contribute to PD.

## Introduction

Parkinson’s disease (PD) is a central nervous system disorder which primarily affects motor skills and oftentimes cognition (Olanow et al., 2009). It is the second most common neurodegenerative condition after Alzheimer’s disease, with an estimated 6 million people affected globally; the disease burden amounted to approximately 700,000 years lost to disability (YLDs) in 2015 (GBD Collaborators, 2016). It is most common in people over 60 years old, and males are affected approximately twice as often as females (Cerri et al., 2019).

Loss of dopaminergic neurons in the substantia nigra is a well characterised feature of PD, and is pathogenic of motor symptoms such as bradykinesia (Surmeier, 2018). Additionally, PD is associated with dementia in 24-31% of cases (Aarsland et al., 2005). Despite the existence of dementia and other non-motor symptoms, the motor symptoms currently form the basis of diagnostic criteria for PD due to their specificity compared to other neurodegenerative disorders (Tolosa et al., 2021); unfortunately this often results in delayed diagnosis since the disease is typically well-established with the development of motor symptoms. Identifying the disease in the prodromal or early stages would potentially improve treatment outcomes, with multiple biomarkers being investigated (Emamzadeh & Surguchov, 2018).

Currently, there are no neuroimaging markers for diagnosing PD, but a variant of SPECT imaging which uses a radiotracer to measure dopamine transporter levels in the striatum DaTSCAN, can be used to support a diagnosis (Bega et al., 2021) and to differentiate PD from essential tremor (ET) where clinical presentation is ambiguous. Beyond that, measures of electrophysiological activity offer a possible biomarker source for understanding PD mechanisms.

Magnetoencephalography (MEG) is a non-invasive functional neuroimaging technology which measures synchronous neural currents using extremely sensitive magnetometers (Hämäläinen et al., 1993). MEG data has excellent temporal resolution (on the order of milliseconds) and good spatial resolution (<5mm with appropriate source modelling). Advanced spatial filtering algorithms have enabled a whole-brain scanning paradigm for MEG analysis, where source activity is estimated in multiple locations of interest simultaneously, providing detailed characterisation of spontaneous neural oscillations and functional connectivity between brain regions. MEG has been used to investigate the electrophysiological correlates of Parkinson’s disease (Boon et al., 2019) showing changes in the spectral content of neural oscillatory activity and functional connectivity. Beta band oscillations (13-30 Hz) are of particular interest in PD due to their role in motor planning and action in the sensorimotor areas of the cerebral cortex (Jurkiewicz et al., 2006). Motor cortical beta band power varied with disease state; greater spontaneous beta band power has been observed in early stage PD patients (Pollok et al., 2012), while reduced power is seen in late stage patients (Heinrichs-Graham et al., 2014).

The concept of “neural slowing”, a blanket term for a shift in the overall frequency content of neural oscillations toward lower frequencies, is commonly employed in neuroimaging studies. Neural slowing is a consistent phenomenology of neurodegenerative conditions including Alzheimer’s disease (Jeong, 2004). However, slowing has also been observed in PD without dementia (Stoffers et al., 2007), suggesting it is not a dementia-specific effect. The underlying neural mechanisms of neural slowing are not clearly understood, but in AD, are hypothesised to stem from neural damage and degeneration, as well as a disrupted balance of excitation/inhibition (E/I) (van Nifterick et al., 2022), with the degree of slowing correlating with amyloid burden and cognitive impairment (A. Wiesman et al., 2022).

In this study, we sought to identify distinct components of neural oscillatory activity which correlate with PD. Firstly, using MEG and phenotype data from the open OMEGA repository, we modelled the general effect of PD on neural oscillatory activity and functional connectivity while controlling for confounding variables such as age, sex, and brain volume. Secondly, using a Partial Least Squares (PLS)-based classifier, we investigated whether oscillatory activity and functional connectivity could accurately predict PD status. We used our PLS-based cross-decomposition to uncover multiple statistically independent patterns of neuropathophysiology within the context of neural slowing. Ultimately, a detailed understanding of oscillopathies in PD could be used with AI-ML to support a cost-effective means of diagnosis and offer targets for personalised treatment strategies for PD.

## Methods

### Participant selection

Participants were selected from the Open MEG Archive (OMEGA) repository, an open database comprising neuroimaging and phenotype data from two separate cohorts: the Pre-symptomatic Evaluation of Experimental or Novel Treatments for Alzheimer Disease (PREVENT-AD) study and QPN (Quebec Parkinson Network). The PREVENT-AD cohort contains only healthy controls, while the QPN scans are a mixture of patients with PD and age-matched healthy controls. Neuroimaging data included resting state MEG scans acquired from a CTF-275 MEG scanner (CTF MEG Neuro Innovations Inc. Coquitlam, BC, Canada) and structural magnetic resonance imaging (MRI) images recorded from a 3T Siemens Magnetom TrioTim (PREVENT-AD) or Prisma-Fit (QPN) data (Siemens Healthcare, etc.). The resting states were acquired with eyes open and had a minimum duration of 5 minutes. Phenotype data included age and sex, and a binary disease status (either control or PD); the PD cohort has been previously studied in (A. I. Wiesman et al., 2023); the authors state that patients “*with mild to moderate (Hoehn and Yahr scale: 1 – 3) idiopathic PD were enrolled* … *all participants had been prescribed a stable dosage of antiparkinsonian medication response prior to study enrollment. Patients were instructed to take their medication as prescribed before research visits, and thus all data were collected in the practically-defined “ON” state.”* All participants had given prior informed consent for the study; access to OMEGA was approved by the ethics board of Hospital for Sick Children (Toronto, CA) and granted under the terms of the OMEGA researcher agreement.

For the regression part of the study, we used data from all participants. A sex-balanced subset of the data were selected to perform classification; see later section. Exclusion criteria included the following: age under 50 years, neurological conditions other than PD, absent MEG or MRI, MEG did not pass quality control. A total of 199 participants were included in the study; their characteristics are shown in **Table 1**.

**Table 1:** Participant characteristics.

| Group | PD | Control |
| --- | --- | --- |
| N | 78 | 121 |
| N(female) | 21 | 89 |
| Mean age (years) | 63 ± 10 | 64 ± 10 |

### MEG data acquisition and preprocessing

MEG data were recorded from the CTF-275 device in the upright position with eyes open at a sampling rate of 2400 Hz with a hardware low pass filter of 1200 Hz applied. Synthetic third-order gradiometry using the CTF-275 reference magnetometer array was used to attenuate environmental magnetic interference. The outline of each participant’s scalp surface was measured using a Polhemus digitizer, which was used to coregister the source model to the sensor coordinate system (see section “Source localisation with beamforming”).

Preprocessing of MEG scans was performed using an automated pipeline built using the open source MNE-Python software package (Gramfort et al., 2013). Firstly, recordings were downsampled to 300 Hz and band pass filtered between 1 and 150 Hz; the power line peak at 60 Hz was removed using a notch filter. A 110 Hz high pass filtered version of the recording was also made, which enabled us to detect typically high frequency muscle artefacts. Continuous head movements were reconstructed from fiducial coil fields; we rejected 7 scans in which the head position drifted more than 7mm from the position at the start of the experiment. No systematic difference in head movement was observed between the control and PD group (p=0.20, independent samples T-test).

Filtered data were then segmented into 10.24 second epochs. Any epoch with a peak-to-peak amplitude exceeding 6000 fT or containing muscle activity was considered to contain interference and excluded from further analysis; this procedure ensured muscle artefacts and low-frequency interference did not contaminate the data. Of the remaining good quality epochs, the first 25 were selected to be analysed, corresponding to 256 seconds of data for each participant. From our initial sample of 214 unique participants, 15 were rejected due to head movement and data quality; further analysis used the remaining 199 participants.

Further data cleaning was performed on the remaining epochs using Independent Components Analysis (ICA). We used the “infomax” method (T. W. Lee et al., 1999) implemented in MNE-python with 80 components to separate out neural and non-neural sources. In the majority of participants, clearly identifiable heartbeat and eye blink components were recovered by ICA; these components were automatically tagged using a proprietary classification algorithm and projected out of the data.

### MRI data acquisition and forward modelling

Each participant’s T1-weighted structural MRI data to generate an individualised, single-layer boundary element model (Mosher et al., 1999) of the inner skull surface using Freesurfer (Fischl, 2012), comprising motion correction and template registration. For each conformed T1-weighted image, we iteratively computed an affine registration from the subject-specific coordinate system to the MNI-152 template space using a robust registration algorithm (Reuter et al., 2010). Using this affine registration, a precomputed template mesh was warped to represent the inner skull boundary. We also derived an approximate cranial vault volume in mm^3 from this mesh for use as a covariate in our contrast model. Finally, seed locations were transformed from MNI to subject coordinates and three axis-aligned unit dipoles were used to calculate three sets of magnetic lead fields.

### Source localization with beamforming

Source modelling was performed using a LCMV beamformer, an adaptive spatial filter which minimises signal variance at a given location subject to the constraint of unit gain (Veen et al., 1997). We calculated the sensor covariance matrix empirically from the cleaned data epochs, applying a regularisation of 5% of the maximum singular value to ensure a stable inversion. Combining this with the lead field information enabled us to calculate optimal beamformer weights. We calculated these LCMV beamformer weights for each cartesian dipole at 78 atlas locations (Automated Anatomical Labelling, AAL – see (Gong et al., 2009)) and 44 network locations. For each seed location, the three cartesian components of source activity were reduced with PCA to a single component, generating a total of 122 source-resolved time series. Each of these time series was then scaled to unit standard deviation.

### Regional spectral power and functional connectivity

We estimated power spectral density (PSDs) from time series for each AAL location using Welch’s method. FFTs were computed on non-overlapping segments of 4096 samples each and then averaged to provide an estimate of the relative frequency contribution of power at each virtual electrode. The resulting power spectra were divided into five a priori frequency bands with a 1 Hz gap to avoid overlap (see **Table 2**). The mean amplitude for each frequency band was then computed, yielding five estimates of band-limited relative power at each AAL node.

**Table 2:** Frequency band definitions.

| Band | Delta | Theta | Alpha | Beta | Gamma |
| --- | --- | --- | --- | --- | --- |
| Frequency (Hz) | 1-3 | 4-8 | 9-12 | 13-29 | 30-45 |

Network node time series were used to estimate connectivity in the same frequency bands. Nodes were selected in the central executive (CEN), default mode (DMN), motor (MOT), visual (VIS), and attention (ATT) networks (see **Supplementary Materials** for node definitions with MNI coordinates). Firstly, the time courses were band-pass filtered for each frequency band. They were then orthogonalized using an iterative symmetric procedure to remove zero-lag correlations, thereby eliminating spurious correlations resulting from magnetic field spread (Colclough et al., 2015). We used the real component of the Hilbert transform to estimate the amplitude envelope for each time series. These envelopes were low pass filtered at 0.5 Hz and downsampled to 1 Hz, and the Pearson correlation coefficient (ρ) between pairs of nodes was calculated, following the procedure outlined in Brookes et al. (2011a). This approach to estimating functional network activity is termed Amplitude Envelope Correlation (AEC).

### Modelling with Contrast of Parameter Estimates

To assess the neurophysiological effects of PD, we modelled spectral and connectivity data *Y* as a linear combination of explanatory phenotype variables *X*. This method is commonly applied when analysing functional MRI experiments (Friston et al., 1994) and has also been applied to MEG data (Quinn et al., 2024) (Gohil et al., 2022). We used a general linear model:

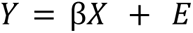

with columns of the design matrix *X* representing disease status, age, sex and head volume, while regional power and connectivity values were vectorised and concatenated to form the matrix of responses *Y*. Fitting the GLM using ordinary least squares resulted in an array of parameter estimates β. The contrast β(*PD*) − β(*control*) was calculated and divided by the regularised (σ → σ + 0. 02 * *max*(σ)) residual variance to compute a pseudo-T-statistic for each MEG feature.

Statistical significance was assessed separately for power and connectivity through permutation testing, using *max*(*abs*(*T*)) as the test statistic; this enables us to avoid a mass-univariate approach at the cost of spatial specificity. We estimated the null distribution of T values by repeatedly shuffling the labels in the contrasted columns and recomputing the GLM 10,000 times. We determined the p-value as the proportion of null statistics more extreme than our observed statistic. A Bonferroni correction factor was applied to control for family-wise errors across separate frequency bands; all p-values for significance tests repeated across bands are reported post-correction, having been multiplied by 5 unless otherwise specified

### Predictive modelling with Partial Least Squares

Partial Least Squares is a cross-decomposition technique that can be applied to regression problems. Given matrices of data observations *X* and *Y*, PLS is used to find linear combinations of features in *X* that relate to linear combinations of features in *Y* and vice versa (Wegelin, 2000). Unlike PCA, PLS is sensitive to low-variance directions in *X* which are nonetheless highly predictive of *Y*.

PLS models have one hyperparameter: the number of latent factors *l* in the joint matrix decomposition of *X* and *Y*. In a multivariate partial least squares scenario with *n* observations, *m* independent variables, and *p* dependent variables, we have two models which are jointly optimised:

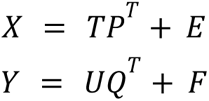

where *X* is the *n* × *m* matrix of independent variables and *Y* is the *n* × *p* matrix of dependent variables. *T* and *U* are *n* × *l* matrices which are projections of *X* and *Y* respectively into latent variables, often called “scores”. *P* (*m* × *l*) and *Q* (*n* × *l*) are loading matrices while *E* and *F* represent intrinsic error.

In this work, PLS is used as a predictive model (Abdi & Williams, 2013), where the MEG metrics are treated as the design matrix *X* and the disease condition is *Y*. The model estimates weightings of the observed MEG metrics which optimally explain the disease condition; this is elaborated in the section titled Classification using PLS models.

### Validating PLS classifier performance

To ensure our classifier was not biased by participant sex, we selected a single stratified subsample of data to perform all classifier training and testing, consisting of 88 total samples with equal numbers of male and female participants in the control and case groups. For all our investigations of performance, we use 40 randomised shuffle splits for cross validation, where 75% of each split is used to train the model and the other 25% is used for testing.

In the case of 2 prediction classes (in this case PD and control), PLS regression can be straightforwardly converted to a classification by converting categorical labels and thresholding the continuous response variable, an approach known as discriminant analysis (L. C. Lee et al., 2018). This enables us to use typical metrics of classifier performance such as the receiver operating characteristic (ROC) curve, which plots true positive rate (TPR) as a function of false positive rate (FPR); to estimate the ROC curve, we vary the decision threshold of the classifier and empirically determine the FPR and TPR for each train-test split of the data. The ROC curve can also be reduced to a single number by calculating the area under the curve (AUC-ROC), which is a balanced summary of classifier performance (accuracy) and ranges from 1 (perfect sensitivity and specificity) to 0.5 (no better than random choice).

The PLS model allows selection of the number of latent components *l* as a hyperparameter. To assess the effect of the number of PLS components on classification performance, we performed a parameter search: for each integer value of *l* in the range [1, 11], a PLS model was trained with *l* components and AUC-ROC was calculated across all validation folds.

To investigate the relative importance of functional connectivity compared to regional oscillatory power, we repeated the above analyses for versions of the models trained with regional power only, AEC connection strengths only, and with all features vectorised and concatenated. In order to determine which MEG data features were important for classification, we retrained the best performing model with all input data. This allows the fitted weights to be used as a proxy for relative feature importance.

## Results

### PD is characterised by topologically diverse cross-spectrum oscillatory changes

Contrasts between control and PD are shown in **Figure 1**. A posterior-dominant pattern of ‘neural slowing’ was observed, with theta band hypersynchronization in PD that was highly significant (p < 0.00001), as well as a spatially similar concomitant decrease in gamma-band activity (p < 0.00001) - in other words, an increase in low frequency activity and a decrease in high frequency activity. Beta band effects were topographically complex, with a significant (p < 0.0005) decrease in the occipital regions accompanied by moderate increases in fronto-temporal regions. Interestingly, we observed a decrease in very slow-wave delta activity (p = 0.002) in fronto-parietal and somatomotor regions. Spatially diffuse alpha-band activity was not significant. T-statistic maps labelled with individual atlas nodes can be found in **Supplementary Figure 1**. The effects of age, sex, and head volume covariates are shown in **Supplementary Figures 2** and **3** - and revealed large effects on oscillatory activity, highlighting the importance of taking such factors into account when analysing spontaneous function.

**Figure 1:**
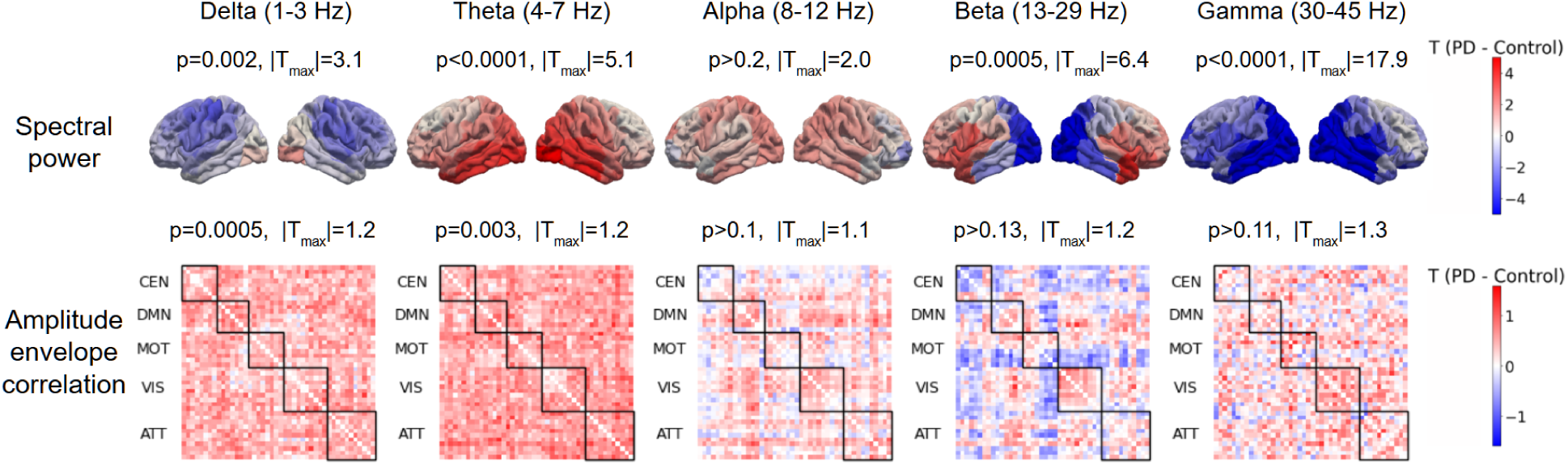
Results of PLS regression. T-statistic images for the contrast of (PD - control) are shown for spectral power (first row) and amplitude envelope correlation (second row). Each column in the overall figure indexes a particular frequency band of interest. In the first row of images, the contrast at each of the 78 AAL nodes is projected onto a schematic image of a brain; left and right lateral orientations are shown. In the second row, AEC contrast values for all network edges are displayed as a matrix where row i and column j index network nodes, and the element M_ij_ is the AEC computed between those nodes. Node groupings corresponding to network definitions (CEN, DMN etc.) are outlined in black. Note the separate colour scales for power and connectivity.

Functional connectivity showed significant global increases across low-frequency coupling, including, delta (p = 0.0005), theta (p = 0.003), with no apparent network specificity; alpha band network activity showed no significant change (p = 0.1). Interestingly, beta band connectivity was not significantly altered in PD (p = 0.23 uncorrected), but showed differential variations in coupling with slight increases within default mode and visual networks, and dysconnectivity between the motor network. Gamma connectivity was not significant (p - 0.12 uncorrected).

### Cortical oscillatory activity reliably predicts PD and with greater accuracy than functional connectivity

**Figure 2** shows the overall performance of the PLS-based linear classifiers; ROC curves for the optimal PLS classifier are shown on the left, while the effect of the number of PLS components on classifier performance is shown on the right. Models trained with regional oscillatory activity performed consistently better than those using connectivity or combined oscillatory & connectivity features (maximum power AUC: 0.87 ± 0.04, network AUC: 0.68 ± 0.04, combined AUC: 0.75 ± 0.04). For the regional oscillatory activity models, adding components initially improves performance, with a clear peak around *l* = 4 components (AUC-ROC: 0.87 ± 0.04); adding subsequent higher dimensional components decreased performance. The combined effect of components 2-4 leads to an increase in AUC-ROC of 0.15 ± 0.04 over and above the performance of a classifier using only one component. A plateau behaviour was observed at *l* = 4 in the combined classifier, suggesting performance was mostly driven by regional power features, while connectivity classifiers appeared unaffected by dimensionality.

**Figure 2:**
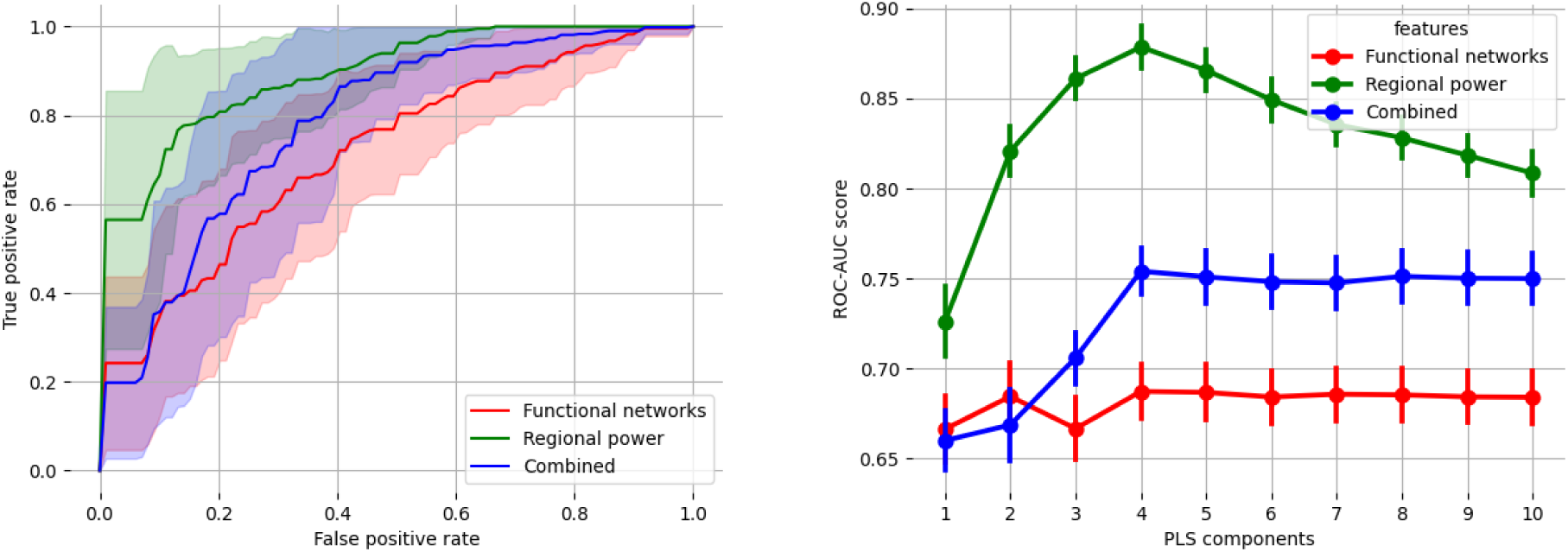
Classification accuracy with PLS is best achieved using cortical oscillatory activity. Left: ROC curves for disease classifiers using different MEG features. Solid lines show the mean ROC across all validation folds; shaded areas show one standard deviation. Four PLS components are used in all cases. Right: cross-validated AUC-ROC scores as a function of PLS components. Vertical bars show standard error on the mean, computed across folds.

### Spatially resolved spectral components independently predict PD

**Figure 3** shows the four discrete components of the optimal power-based PLS classifier when trained on all input data; weights were premultiplied by 1 or -1 such that their correlation with the target variable is always positive. The *X* weightings for each component are displayed in the left hand plot, while the *X* scores for each component are shown on the combined jitter and box plots on the right hand side. As expected, the most discriminative component (*l* = 1) is very similar to the observed PD > control contrast effect, showing posterior theta slowing effectively differentiates controls from patients. Component 2 shows broad decreases in delta and alpha frequencies, while frontal midline theta appears to decrease; this is accompanied by increases in beta and gamma band activity, suggesting an overall shift to higher frequencies. Component 3 shows a global slowing effect with delta showing the largest effect, while component 4 appears to emphasise a global theta decrease accompanied by bilaterally increased beta over superior temporal areas.

**Figure 3:**
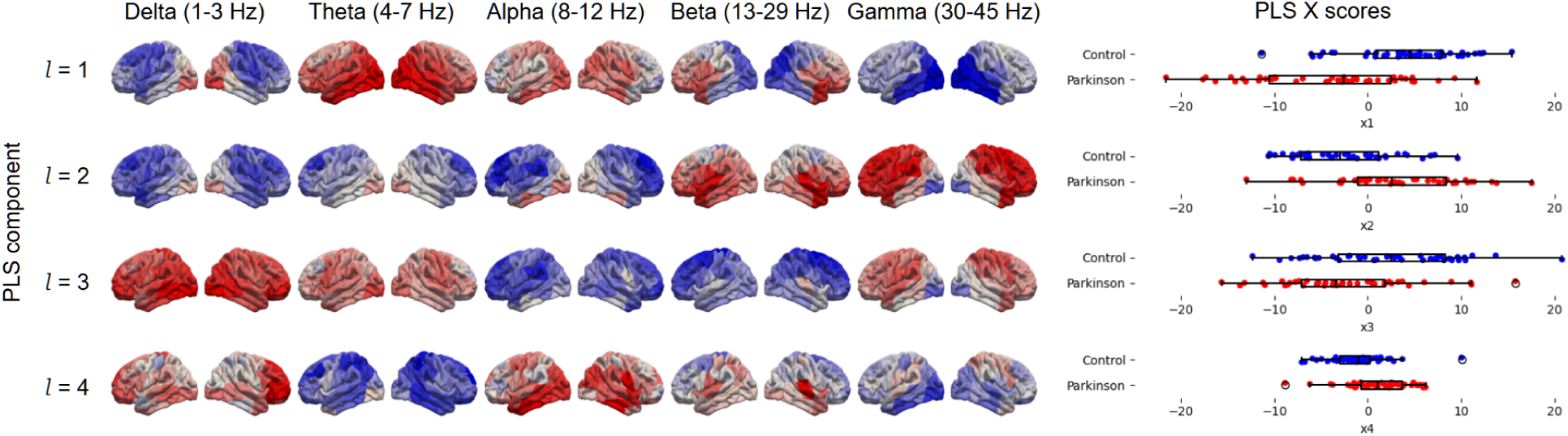
Distinct oscillatory activity features of the optimal PLS classification model. The left subfigure displays PLS weights from the best performing model with *l* = 4 components, trained on power features only. The right subfigure shows the distribution of the four orthogonal model scores across the control and case group. Each row indexes a single PLS component, while each column indexes the frequency band of interest. Excess power in positively weighted regions (red) increases the likelihood of being classified as PD, while excess power in negatively weighted regions (blue) decreases this likelihood.

## Discussion

### Summary

Neurodegenerative processes in Parkinson’s disease (PD) have mechanistic and measurable effects on neural oscillatory activity in the brain. Using a contrast-based modelling approach, we revealed the neurophysiological effects of PD, while controlling for the confounding effects of age, sex and head size. We found that each of these variables were associated with significant effects on brain activity, demonstrating the need for effective confound modelling. Additionally, we used a PLS-based predictive model to classify the disease status of participants with competitive sensitivity and specificity, achieving an AUC-ROC of 0.87 ± 0.04, broadly characterised as “good” or “very good” (Hond et al., 2022). Interpretation of the model weights reveals four linear combinations of oscillatory features, each of which independently predicts PD. These components imply a diverse set of spatially and spectrally distinct oscillatory changes across the cortical surface. The observed correspondence between classifier weights and group-level contrast suggests that the classifier is leveraging genuine disease-associated neural differences (rather than incidental biases in samples), and applying cross-validation demonstrates that the classifier generalises to new samples.

### Oscillatory slowing in PD

The effects on Parkinson’s disease on neural oscillations have been studied for over 50 years (Neufeld et al., 1988). A recent review of PD-related changes in MEG data (Boon et al., 2019) reports a slew of alterations in electrical activity associated with PD. These changes appear to vary depending on the disease stage, but one robust result is an increase in low-frequency oscillations, observed in both early and late stage PD patients with respect to controls. Early EEG studies of PD (Soikkeli et al., 1991) and dementia in general (Hughes et al., 1989) noted significant slowing in both groups. Oscillatory slowing in PD has been observed with MEG (A. I. Wiesman et al., 2023) using an analysis pipeline based on Brainstorm (Tadel et al., 2011), suggesting that these changes are robust to choice of analysis tool.

In the current study, we observed multiple distinct modes of oscillatory change which were directly predictive of PD, which could be interpreted as separate disease processes. At least two of these components (1 and 3) are diffuse and unambiguously identified with neural slowing. However, these components differ greatly in their spatial distribution and most predictive frequency bands: component 1 exhibits a pattern of theta-dominant slowing with a posterior-oriented gradient typically identified with PD, while component 3 is dominated by a global diffuse delta-band effect. Components 2 and 4 are more difficult to describe in terms of slowing, with component 2 showing an overall shift to higher frequencies, and component 4 showing details such as bilaterally increased beta power over superior temporal lobes.

Capturing these fine spatial details relies on the specificity of source-resolved MEG data using MRI-derived individual head models. Our results suggest that neural slowing – while a useful summary measure for determining the overall magnitude and direction of spectral changes – may fail to capture important details of spectral effects, especially with regards to the observed differences in theta- and delta-wave slowing in PD.

### Predicting PD from MEG

EEG and MEG have been previously leveraged as predictive biomarkers for PD and PD dementia (PDD); for example, quantitative EEG has been used to predict development of dementia in PD, with an increased hazard ratio (HR = 3.0; p = 0.004) for median theta band power (Klassen et al., 2011). We found competitive performance (AUC-ROC: 0.87 ± 0.04) using our PLS model, gaining the best performance using only spectral power features and a relatively low number of linear components (l = 4) despite the inherently high dimensionality of the MEG data. Our PLS classifier identified features previously determined to be highly predictive of PDD conversion, such as lowered beta power (Olde Dubbelink et al., 2014) and higher delta/theta power (Caviness et al., 2015). Previously published studies on classifying PD from M/EEG report a range of accuracy values: 82% using Lempel-Ziv complexity (Gómez et al., 2011), 88% using a deep learning approach (Oh et al., 2020), and 93% using a linear predictive coding approach (Anjum et al., 2020). As PLS is a fundamentally linear method, its raw predictive performance given unlimited training data will be strictly inferior in comparison with nonlinear “deep learning” approaches, such as neural network or transformer architectures. However, PLS retains some key advantages in the context of our dataset and goals. Firstly, PLS is ideally suited to our small neuroimaging dataset with more features than observations, providing practical classification accuracy with a fraction of the sample size required to train a deep learning classifier (often in the tens or hundreds of thousands of samples). PLS works with the many correlated MEG features without a regularisation hyperparameter (as with e.g. ridge regression), which makes cross-validation easier. Lastly, PLS facilitates interpretation of the features directly relevant to classification, whereas deep learning methods often present a “black box”; as such, this coincides with our goal of understanding neurophysiological differences.

We found that our best performing model uses only oscillatory features, excluding functional network information completely. Given the proven utility of the functional network paradigm in neuroimaging, this result is somewhat surprising; however, previous studies using MEG for biomarker discovery have reported similar results in which source power outperformed connectivity metrics in predictive power (Engemann et al., 2020). A penalty to performance is paid when including more features, which is a consequence of regularisation applied to prevent overfitting; thus, including more features must be justified by the predictive power of those features. The lower performance of network features might therefore be partially explained by their quantity (5120 network edges vs. 390 power nodes); an approach using fewer and more judiciously chosen seed locations might result in better performance.

### Limitations

Our study was subject to several limitations, both in terms of the available data and the analysis which was performed on that data. One such limitation of our available data was the encoding of disease status as a simple binary variable. While this simplifies the analysis, significant variation in Parkinsonian symptoms is elided. It is known that disease stage and symptom severity, medication status and presence of tremor or dyskinesia also have impacts on the observed electrophysiological data, and this was also not investigated. In general, we were able to use the large sample size (*N* = 199) to increase statistical power in the group contrast and effectively cross-validate the classifier results; however, the requirement for sex-balanced input data for the classifier led to a reduction in the available data for training and evaluation. After excluding non-PD neurological diagnoses, the limiting group was females with PD (*N* = 21); this is partially a consequence of the higher prevalence of PD in males (Cerri et al., 2019). Nevertheless, the observed significant effects of head volume and sex make it hard to justify the use of unbalanced data for classification. Recording more data from women with PD would greatly increase the available sample size.

Our MEG analysis pipeline, while keeping to established guidelines (Gross et al., 2013) as much as possible, has some fundamental limitations. For instance, the scaling of the virtual electrode time series to unit standard deviation is a common choice in resting state MEG analysis. However, it has the effect that information about the absolute signal magnitude in discrete frequency bands is lost: instead, the spectral values presented should be interpreted as fractional or relative weightings of frequency content.

## Conclusion

Our findings suggest that neural slowing effects in PD can be decomposed into multiple orthogonal components, exhibiting complex patterns of variation across the cortical surface which differ both in spectral content and spatial organisation. While each of these components is independently predictive of PD, the optimal classification is achieved with a combination of these components, reliably distinguishing individuals with PD from matched controls with competitively high sensitivity and specificity in an age- and sex-balanced sample. These results suggest that neural oscillations measured with source-resolved MEG be further investigated as a biomarker for disease staging or early detection in PD and other neurodegenerative disorders which exhibit neural slowing.

## Data Availability

Data are available from the Omega repository.

https://www.mcgill.ca/bic/neuroinformatics/omega

## Supplementary material

**Supplementary Figure 1:**
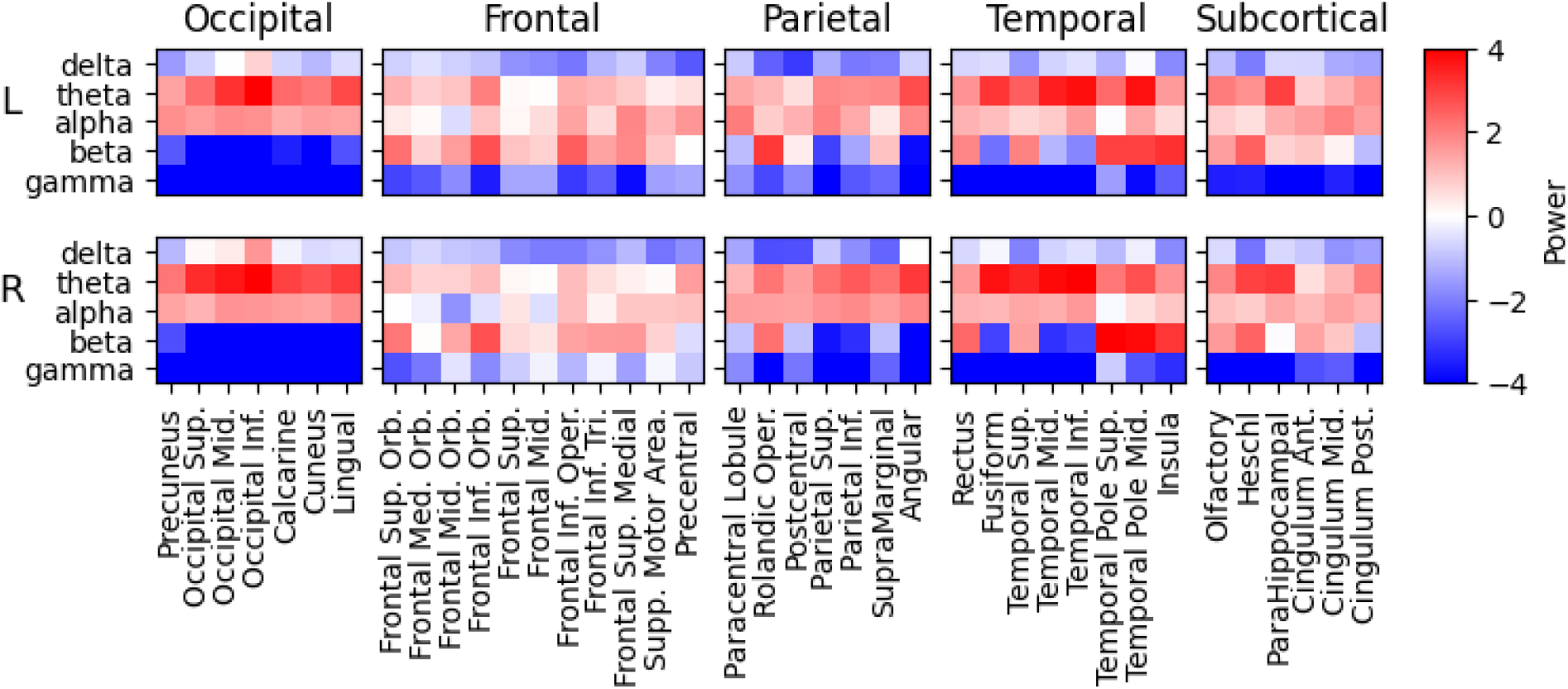
Labelled map of individual T-statistic values for PD > control contrast. Each of the 10 matrices present pseudo-T values for individual brain regions within a lobe grouping; within each image, columns index regions (i.e. AAL nodes) and rows index frequency bands. The images are further arranged into two rows, the upper and lower rows denoting left and right hemisphere regions, respectively. All images use the same global colour map.

**Supplementary Figure 2:**
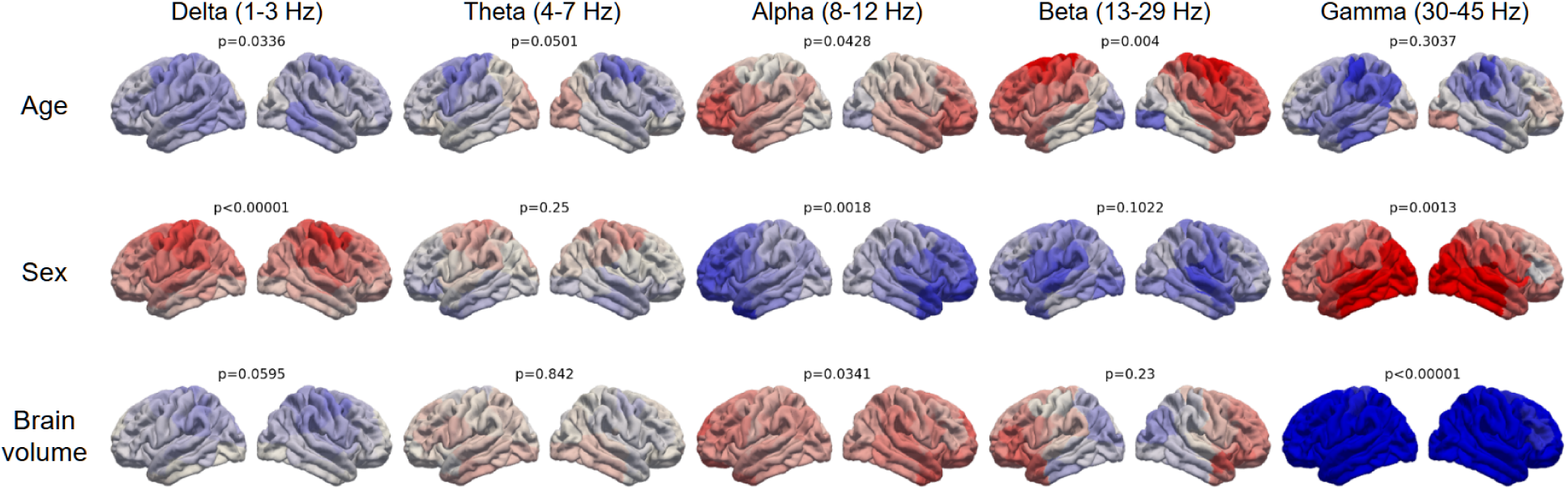
Oscillatory activity contrast maps for age, sex and volume, with p-values uncorrected for FWER. Every factor exhibited large moderating effects on a number of frequency bands.

**Supplementary Figure 3:**
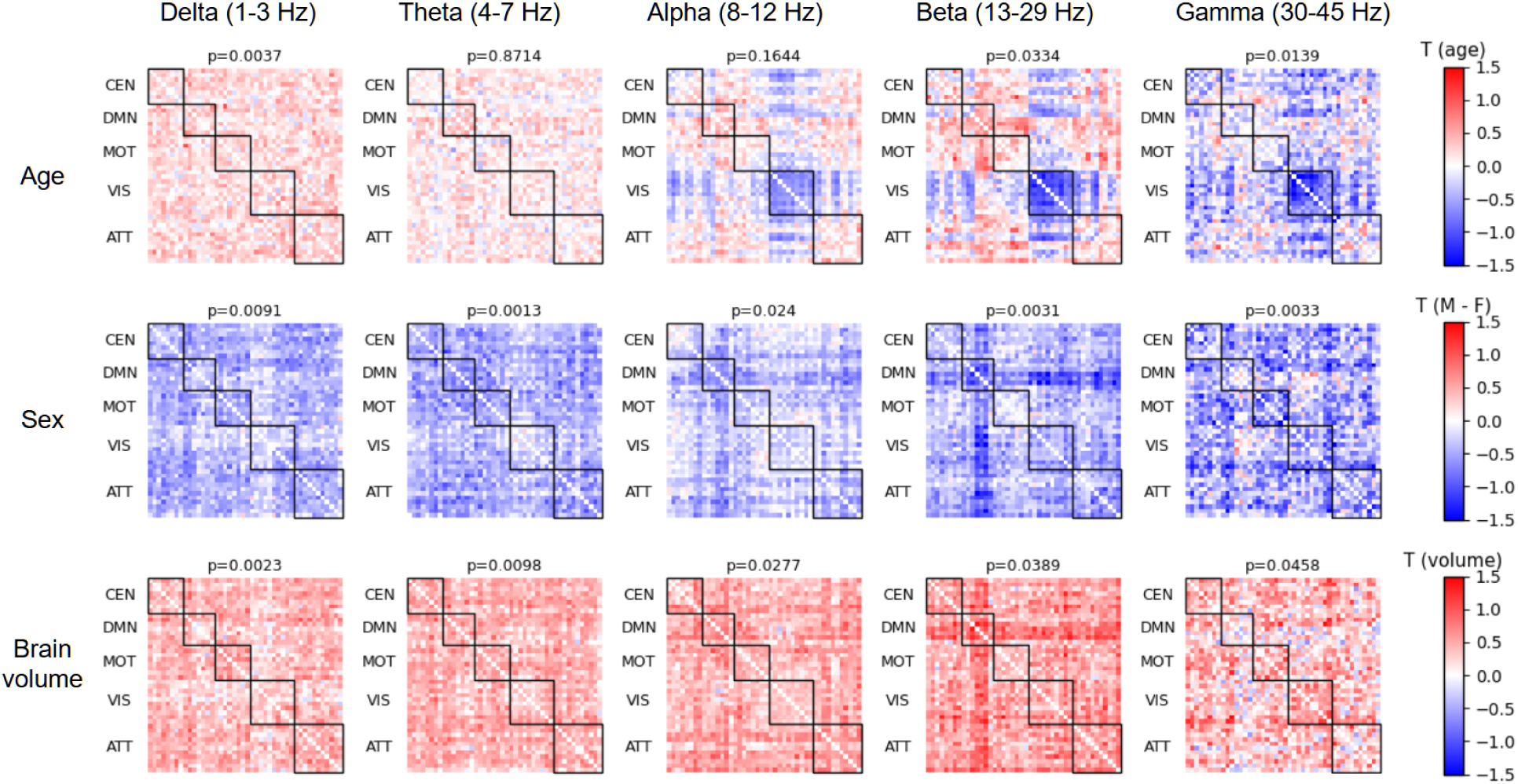
Network contrast maps for age, sex and volume. p-values are uncorrected for FWER.

### Network Nodes Definitions

#### Central Executive Network

RIPS [25, -62, 53]

RVV [36, -62, 0]

LVV [-44, -60, -6]

RSMG [32, -38, 38]

RSLOC [26, -64, 54]

LSLOC [-26, -60, 52]

RFEF [28, -4, 58]

LFEF [-26, -8, 54]

#### Default Mode Network

LAG [-43, -76, 35]

RAG [51, -64, 32]

PCC [-3, -54, 31]

vMPFC [-2, 51, 2]

dMPFC [-13, 52, 23]

RMPFC [2, 53, 24]

LITG [-57, -25, -17]

#### Motor network

Precentral_L [-39.0, -7.0, 50.0]

Precentral_R [41.0, -10.0, 51.0]

Postcentral_L [-43.0, -24.0, 47.0]

Postcentral_R [41.0, -27.0, 51.0]

Parietal_Sup_L [-24.0, -61.0, 58.0]

Parietal_Sup_R [26.0, -60.0, 61.0]

Parietal_Inf_L [-43.0, -47.0, 45.0]

Parietal_Inf_R [46.0, -48.0, 48.0]

#### Attention network

RSMG [52, -48, 28]

RFEF [30, -13, 53]

LFEF [-26, -12, 53]

LpIPS [-25, -67, 48]

RpIPS [23, -69, 49]

LMT [-43, -72, -8]

RMT [42, -70, -11]

RMFG [41, 17, 31]

RPCS [41, 2, 50]

RSTG [58, -48, 10]

RVFC [40, 21, -4]

#### Visual network

LV1 [-3, -101, -1]

RV1 [11, -88, -4]

LV2d [-8, -99, 7]

RV2d [14, -96, 13]

LV3 [-9, -96, 13]

RV3 [20, -95, 18]

LV4 [-31, -77, -17]

RV4 [27, -71, -14]

LV7 [-23, -78, 26]

RV7 [32, -78, 25]

#### Abbreviations

“RIPS”: “Right Intra Parietal Sulcus”,

“RVV”: “Right Ventral Visual”,

“LVV”: “Left Ventral Visual”,

“RSMG”: “Right Supramarginal Gyrus”,

“RSLOC”: “Right Superior Lateral Occipital Cortex”,

“LSLOC”: “Left Superior Lateral Occipital Cortex”,

“RFEF”: “Right Frontal Eye Field”,

“LFEF”: “Left Frontal Eye Field”,

“LAG”: “Left Angular Gyrus”,

“RAG”: “Right Angular Gyrus”,

“PCC”: “Posterior Cingulate Cortex”,

“vMPFC”: “Ventromedial Prefrontal Cortex”,

“dMPFC”: “Dorsomedial Prefrontal Cortex”,

“RMPFC”: “Rostral Medial Prefrontal Cortex”,

“LITG”: “Left Inferior Temporal Gyrus”,

“Precentral_L”: “Precentral Left”,

“Precentral_R”: “Precentral Right”,

“Postcentral_L”: “Postcentral Left”,

“Postcentral_R”: “Postcentral Right”,

“Parietal_Sup_L”: “Parietal Superior Left”,

“Parietal_Sup_R”: “Parietal Superior Right”,

“Parietal_Inf_L”: “Parietal Inferior Left”,

“Parietal_Inf_R”: “Parietal Inferior Right”,

“LV1”: “Left Visual 1”,

“RV1”: “Right Visual 1”,

“LV2d”: “Left Visual 2 (Dorsal)”,

“RV2d”: “Right Visual 2 (Dorsal)”,

“LV3”: “Left Visual 3”,

“RV3”: “Right Visual 3”,

“LV4”: “Left Visual 4”,

“RV4”: “Right Visual 4”,

“LV7”: “Left Visual 7”,

“RV7”: “Right Visual 7”,

“LpIPS”: “Left posterior Intra-Parietal Sulcus”,

“RpIPS”: “Right posterior Intra-Parietal Sulcus”,

“LMT”: “Left Middle Temporal”,

“RMT”: “Right Middle Temporal”,

“RMFG”: “Right Middle Frontal Gyrus”,

“RPCS”: “Right Precentral Sulcus”,

“RSTG”: “Right Superior Temporal Gyrus”,

“RVFC”: “Right Ventro-Frontal Cortex”

#### AAL nodes with MNI coordinates

Rectus_L: [ -5. 36. -20.]

Olfactory_L: [ -8. 14. -13.]

Frontal_Sup_Orb_L: [-17. 46. -15.]

Frontal_Med_Orb_L: [-5. 53. -9.]

Frontal_Mid_Orb_L: [-31. 49. -11.]

Frontal_Inf_Orb_L: [-36. 29. -13.]

Frontal_Sup_L: [-19. 33. 41.]

Frontal_Mid_L: [-34. 31. 34.]

Frontal_Inf_Oper_L: [-49. 11. 18.]

Frontal_Inf_Tri_L: [-46. 29. 13.]

Frontal_Sup_Medial_L: [-5. 48. 30.]

Supp_Motor_Area_L: [-6. 4. 60.]

Paracentral_Lobule_L: [ -8. -27. 69.]

Precentral_L: [-39. -7. 50.]

Rolandic_Oper_L: [-47. -10. 13.]

Postcentral_L: [-43. -24. 47.]

Parietal_Sup_L: [-24. -61. 58.]

Parietal_Inf_L: [-43. -47. 45.]

SupraMarginal_L: [-56. -35. 29.]

Angular_L: [-44. -62. 34.]

Precuneus_L: [ -8. -57. 47.]

Occipital_Sup_L: [-17. -86. 27.]

Occipital_Mid_L: [-33. -82. 15.]

Occipital_Inf_L: [-36. -80. -9.]

Calcarine_L: [ -7. -80. 5.]

Cuneus_L: [ -6. -81. 26.]

Lingual_L: [-15. -69. -6.]

Fusiform_L: [-31. -41. -22.]

Heschl_L: [-42. -20. 9.]

Temporal_Sup_L: [-53. -22. 6.]

Temporal_Mid_L: [-56. -35. -4.]

Temporal_Inf_L: [-50. -29. -25.]

Temporal_Pole_Sup_L: [-40. 14. -21.]

Temporal_Pole_Mid_L: [-37. 13. -35.]

ParaHippocampal_L: [-21. -17. -22.]

Cingulum_Ant_L: [-4. 34. 13.]

Cingulum_Mid_L: [ -6. -16. 40.]

Cingulum_Post_L: [ -5. -44. 23.]

Insula_L: [-35. 5. 2.]

Rectus_R: [ 8. 34. -19.]

Olfactory_R: [ 10. 15. -13.]

Frontal_Sup_Orb_R: [ 18. 47. -15.]

Frontal_Med_Orb_R: [ 8. 50. -9.]

Frontal_Mid_Orb_R: [ 33. 51. -12.]

Frontal_Inf_Orb_R: [ 41. 31. -13.]

Frontal_Sup_R: [22. 30. 43.]

Frontal_Mid_R: [37. 32. 33.]

Frontal_Inf_Oper_R: [50. 14. 20.]

Frontal_Inf_Tri_R: [50. 29. 13.]

Frontal_Sup_Medial_R: [ 9. 50. 29.]

Supp_Motor_Area_R: [ 8. -1. 61.]

Paracentral_Lobule_R: [ 7. -33. 67.]

Precentral_R: [ 41. -10. 51.]

Rolandic_Oper_R: [52. -8. 13.]

Postcentral_R: [ 41. -27. 51.]

Parietal_Sup_R: [ 26. -60. 61.]

Parietal_Inf_R: [ 46. -48. 48.]

SupraMarginal_R: [ 57. -33. 33.]

Angular_R: [ 45. -61. 37.]

Precuneus_R: [ 10. -57. 42.]

Occipital_Sup_R: [ 24. -82. 29.]

Occipital_Mid_R: [ 37. -81. 18.]

Occipital_Inf_R: [ 38. -83. -9.]

Calcarine_R: [ 16. -74. 8.]

Cuneus_R: [ 13. -81. 27.]

Lingual_R: [ 16. -68. -5.]

Fusiform_R: [ 34. -40. -22.]

Heschl_R: [ 46. -18. 9.]

Temporal_Sup_R: [ 58. -23. 5.]

Temporal_Mid_R: [ 57. -39. -3.]

Temporal_Inf_R: [ 53. -32. -24.]

Temporal_Pole_Sup_R: [ 48. 13. -18.]

Temporal_Pole_Mid_R: [ 44. 13. -34.]

ParaHippocampal_R: [ 25. -16. -22.]

Cingulum_Ant_R: [ 8. 36. 14.]

Cingulum_Mid_R: [ 8. -10. 38.]

Cingulum_Post_R: [ 7. -43. 20.]

Insula_R: [39. 5. 1.]

